# Beyond Access: Racial Differences in Income-Related Gains in Tooth Retention by Dental Care Context

**DOI:** 10.64898/2026.03.25.26349348

**Authors:** Kym M. McCormick

## Abstract

**Objectives:** To test whether the association between household income and tooth retention differs by race/ethnicity and whether this interaction varies by reason for the most recent dental visit among US adults.

**Methods:** We analyzed 13,190 adults in the National Health and Nutrition Examination Survey (2009–2018). Survey-weighted linear regression estimated interactions between household income and race/ethnicity in models of tooth retention, stratified by reason for last dental visit.

**Results:** Higher income was associated with greater tooth retention across groups, but income-related gains were larger for Non-Hispanic White adults than for Non-Hispanic Black and Mexican American adults, particularly in problem-focused care settings. In problem-focused visits, each higher income category was associated with 0.5 additional teeth among White adults (95% CI = 0.4, 0.6) versus 0.2 (95% CI = 0.0, 0.4) among Black adults and 0.1 (95% CI = −0.1, 0.3) among Mexican American adults. Racial differences were attenuated in routine check-up contexts.

**Conclusions:** Income-related gains in tooth retention differed by race/ethnicity and dental care context.

**Public Health Implications:** Expanding access alone may be insufficient to reduce racial inequities in oral health.

## Introduction

Tooth loss reflects the cumulative effects of oral disease, treatment decisions, and patterns of dental care use across the life course and is associated with impaired nutrition, social participation, and quality of life.^1^ Although oral health in the United States has improved overall, substantial racial and ethnic disparities in tooth retention persist.^2^ These disparities are commonly attributed to differences in socioeconomic position and access to dental care, including dental insurance coverage and patterns of service use, factors that are often assumed to narrow gaps when improved.^2,3^

However, racial disparities remain evident even among individuals with similar income, educational attainment, and access to dental services.^4^ This pattern suggests that socioeconomic resources do not translate into oral health benefits equally across racial groups. Consistent with the concept of “diminished returns,” racial and ethnic minority groups often experience smaller health gains from equivalent socioeconomic resources.^5^ This literature points to structural racism as an important mechanism shaping not only access to resources, but also how effectively those resources are converted into health within clinical and institutional settings.^6^

Dental care provides a particularly relevant context in which to examine these processes. The U.S. dental system is highly privatized, and treatment decisions frequently involve discretion between preventive, restorative, and extraction-oriented care.^7^ In this context, the reason for a dental visit—whether for a routine check-up or for a specific problem—captures an important dimension of clinical care. Preventive visits emphasize continuity and early intervention and may help to reduce accumulated disadvantage, whereas problem-focused visits reflect more reactive and episodic engagement with the dental system and may amplify disadvantage through delayed intervention and extraction-oriented treatment.^8^ Income and race may therefore interact differently across these care contexts.

Using nationally representative NHANES data, we examined whether the association between household income and tooth retention differs by race/ethnicity and whether this interaction varies by reason for last dental visit, adjusting for age, sex, insurance, education, and prior-year dental access. By modeling interactions between income, race/ethnicity, and dental care context, this study applies a diminished-returns framework to oral health. Differences in income-related gains in tooth retention are interpreted as outcome-based indicators of how structural processes may shape the translation of socioeconomic resources into health within dental care systems.

## Methods

### Data Source and Sample

We analyzed five NHANES cycles (2009–2018), a nationally representative multistage survey. Adults aged ≥20 years with a complete oral exam and non-missing covariates were included (N = 13,190). Third molars were excluded (maximum 28 teeth).

### Measures

Outcome: number of retained natural teeth. Variable of interest: self-identified race/ethnicity (Non-Hispanic White, Non-Hispanic Black, Mexican American). Other Hispanic and multiracial respondents were excluded due to small sample sizes; sensitivity analyses were similar. Household income was modeled as ordinal income-to-poverty ratio categories (1–12). The effect modifier was reason for most recent dental visit (check-up vs. problem-focused). Covariates: age, sex, dental insurance, education (college vs. no college), and prior-year dental access. Missingness was <1% for all variables except household income missingness (∼10– 14%). This was addressed via complete-case analysis and may result in conservative estimates of income gradients.

### Statistical Analysis

Survey-weighted linear regression modeled tooth retention by race, income, and their interaction, stratified by visit reason. Predicted marginal means with 95% CIs were generated. Analyses incorporated NHANES weights, strata, and PSUs. Results are reported as unstandardized coefficients (b) with 95% CIs.

## Results

### Sample Characteristics

The sample included 7,019 Non-Hispanic White, 3,973 Non-Hispanic Black, and 2,198 Mexican American adults. The weighted mean number of retained teeth was 23.2 (SE = 0.1). Sample characteristics are presented in Table 1.

**Table 1.** Sample characteristics by racial and ethnic group, NHANES 2009–2018.

| **Characteristic** | **Level** | **Non-Hispanic**<br>**White**<br>**(n = 7,019)** | **Non-Hispanic**<br>**Black**<br>**(n = 3,973)** | **Mexican American**<br>**(n = 2,198)** |
| --- | --- | --- | --- | --- |
| **Dentition** |  |  |  |  |
|  | mean (SE) | 23.37 (0.20) | 22.04 (0.17) | 24.81 (0.19) |
| **Sex** |  |  |  |  |
|  | Male | 3,478 (49.6%) | 1,893 (47.6%) | 1,042 (47.4%) |
|  | Female | 3,541 (50.4%) | 2,080 (52.4%) | 1,156 (52.6%) |
| **Age (years)** |  |  |  |  |
|  | 20–34 | 1,645 (23.4%) | 993 (25%) | 591 (26.9%) |
|  | 35–59 | 2,672 (38.1%) | 1,641 (41.3%) | 974 (44.3%) |
|  | 60+ | 2,702 (38.5%) | 1,339 (33.7%) | 633 (28.8%) |
| **Household Income (tertiles)<sup>a</sup>** |  |  |  |  |
|  | Lower | 2,160 (30.8%) | 1,484 (37.4%) | 753 (34.3%) |
|  | Middle | 2,183 (31.1%) | 1,328 (33.4%) | 886 (40.3%) |
|  | Higher | 2,676 (38.1%) | 1,161 (29.2%) | 559 (25.4%) |
| **Educational attainment<sup>b</sup>** |  |  |  |  |
|  | No college | 2,585 (36.8%) | 1,778 (44.8%) | 1,469 (66.8%) |
|  | College | 4,434 (63.2%) | 2,195 (55.2%) | 729 (33.2%) |
| **Reason for last dental visit<sup>c</sup>** |  |  |  |  |
|  | Check-up | 4,110 (58.6%) | 1,956 (49.2%) | 1,103 (50.2%) |
|  | Problem | 1,855 (26.4%) | 1,358 (34.2%) | 710 (32.3%) |
|  | Other | 1,054 (15%) | 659 (16.6%) | 385 (17.5%) |
| **Access to dental care<sup>d</sup>** |  |  |  |  |
|  | Accessed | 5,705 (81.3%) | 2,926 (73.6%) | 1,584 (72.1%) |
|  | Could not access | 1,314 (18.7%) | 1,047 (26.4%) | 614 (27.9%) |
| **Insurance coverage** |  |  |  |  |
|  | Covered | 6,111 (87.1%) | 3,250 (81.8%) | 1,475 (67.1%) |
|  | Not covered | 908 (12.9%) | 723 (18.2%) | 723 (32.9%) |

Note. Dentition values are weighted means (SE). Other values are unweighted n (column percentage). Percentages may not sum to 100 due to rounding.
<sup>a</sup> Missing: NHW = 510, NHB = 579, MA = 373.
<sup>b</sup> Missing: NHW = 1, NHB = 10, MA = 1.
<sup>c</sup> Missing: NHW = 0, NHB = 0, MA = 0.
<sup>d</sup> Missing: NHW = 0, NHB = 0, MA = 1.
NHANES, National Health and Nutrition Examination Survey; NHW, Non-Hispanic White; NHB, Non-Hispanic Black; MA, Mexican American.

### Race × Income Interactions by Reason for Dental Visit

Higher income was associated with greater tooth retention in both care contexts, but gradients differed markedly by visit reason (Figure 1).

**Figure 1.**
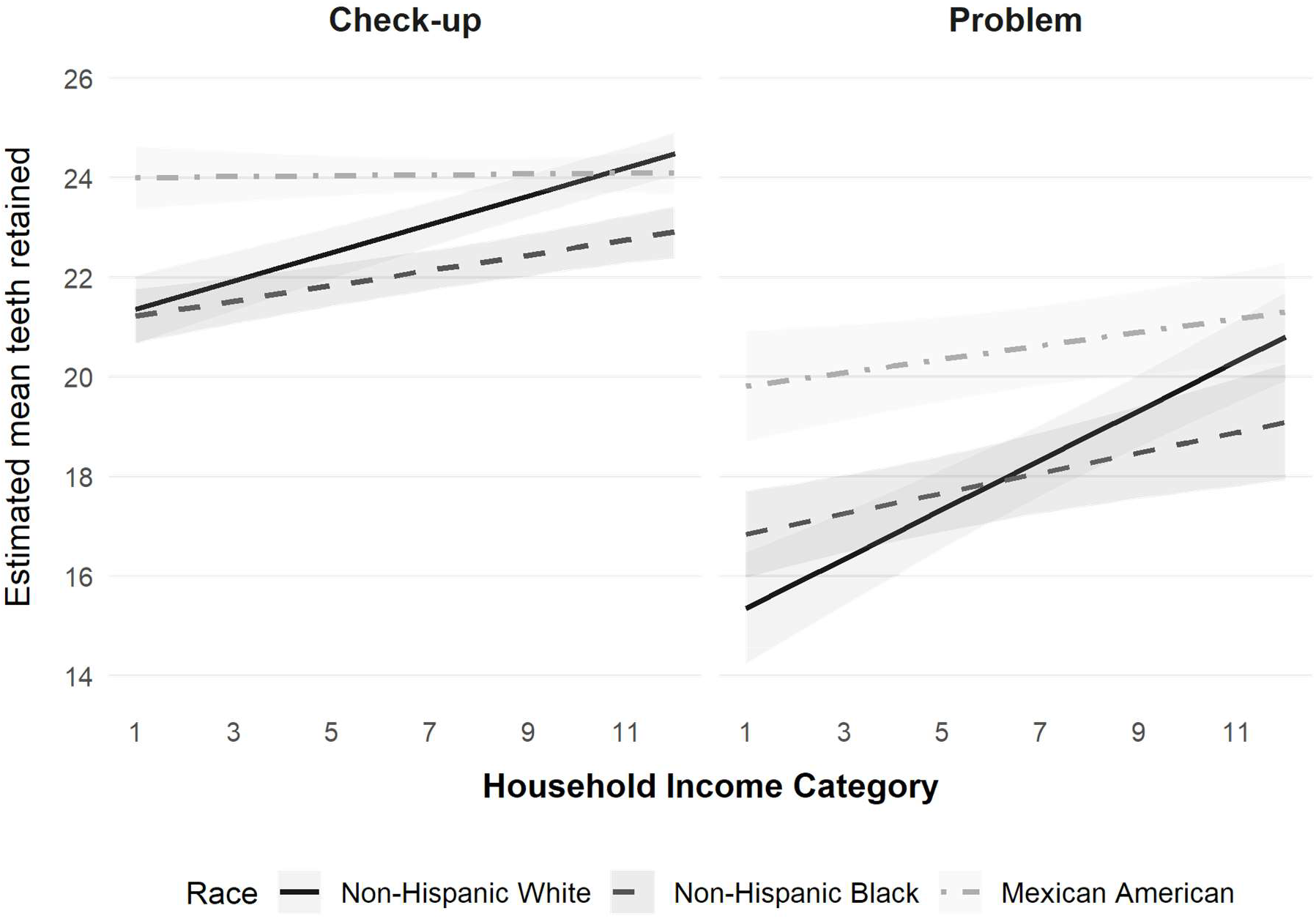
Estimated mean number of natural teeth retained by household income and race, stratified by reason for dental visit. Predicted values are derived from survey-weighted linear regression models including race/ethnicity, household income category (treated as ordinal), and their interaction, adjusted for age (continuous), sex (male vs. female), education attainment (college vs. no college), access to care (access vs. no access), and insurance coverage (covered vs not covered). The left panel represents individuals whose last dental visit was for a routine check-up, while the right panel represents those whose visit was problem-focused. Shaded bands represent 95% confidence intervals. All estimates account for the complex NHANES sampling design and pooled examination weights (2009–2018).

#### Problem-focused visits

The income gradient for Non-Hispanic White adults was nearly twice as steep (b = 0.50 teeth per income category; 95% CI: 0.37, 0.62). Racial differences in gradients were larger (Non-Hispanic Black: b = −0.29; 95% CI: −0.46, −0.12; Mexican American: b = −0.36; 95% CI: −0.53, −0.19). Across the income range, predicted retention increased by ≈5.5 teeth for Non-Hispanic White adults versus ≈2.2 (Non-Hispanic Black) and ≈1.5 (Mexican American). Outcome variability was substantially higher in this stratum.

#### Check-up visits

Income was positively associated with tooth retention for Non-Hispanic White adults (b = 0.28 teeth per income category; 95% CI: 0.22, 0.34). Income gradients were smaller for Non-Hispanic Black (interaction b = −0.13; 95% CI: −0.22, −0.04) and Mexican American adults (b = −0.27; 95% CI: −0.37, −0.18). Predicted tooth retention remained high across income for all groups (≈21–24 teeth), with overlapping confidence intervals.

A three-way race × income × visit-reason interaction indicated income gradients differed across care contexts, with confidence intervals excluding the null for interaction terms involving problem-oriented care.

## Discussion

This study applies a diminished-returns framework to oral health by examining whether income-related gains in tooth retention differ by race/ethnicity and dental care context. Income-related gains were not uniform across racial groups, and the degree of stratification varied by reason for the most recent dental visit. These findings suggest that structural inequality may operate not only through access to dental care, but also within dental care delivery.

The contrast between care contexts is consistent with theoretical accounts of how preventive versus reactive care systems may constrain or amplify accumulated disadvantage.^7^ Preventive care is characterized by continuity, early intervention, and a treatment philosophy oriented toward preservation, conditions that may allow socioeconomic resources to be translated into oral health benefits more equitably. In contrast, problem-focused care is typically episodic and crisis-driven and may involve greater clinical discretion in treatment decisions. Within such contexts, differential treatment pathways, including the balance between extraction- and restoration-oriented care, may contribute to unequal translation of socioeconomic resources into retained dentition.^6^ Although treatment decisions were not directly measured, the observed patterns are consistent with processes operating within clinical care.

Importantly, models adjusted for dental insurance coverage, educational attainment, and prior-year dental access, suggesting that the observed interaction is not fully explained by differences in access to care. This supports an interpretation in which processes occurring within dental care delivery, including continuity of care, treatment decision-making, and the broader structural conditions shaping clinical encounters, may play a role.^5^

Several limitations warrant consideration. The cross-sectional design precludes causal inference, and the observed gradients reflect accumulated lifetime experiences rather than changes over time. Reason for last dental visit was measured at a single time point and may not fully capture long-term care patterns. In addition, the exclusion of other Hispanic and multiracial respondents limits generalizability to those groups.

### Public Health Implications

These findings suggest that expanding access to dental care alone may be insufficient to eliminate racial disparities in tooth retention. Even among adults who engaged with dental services, racial differences in income-related gains persisted in problem-focused care contexts. Policies that promote early and routine preventive dental visits (such as Medicaid coverage of adult preventive services, community dental health programs, and incentive structures that reward tooth-preserving treatment) may help reduce inequities in oral health outcomes. Evaluating dental care systems using outcomes such as tooth retention, rather than utilization alone, may provide a clearer assessment of whether care functions as an equalizer or an amplifier of structural inequality.

## Data Availability

All data produced in the present study are available upon reasonable request to the author

https://www.cdc.gov/nchs/nhanes

## Acknowledgement

The author thanks Dr. Joao Luiz Bastos for suggesting literature that informed the early development of this work.

